# GatorTron: A Large Language Model for Clinical Natural Language Processing

**DOI:** 10.1101/2022.02.27.22271257

**Authors:** Xi Yang, Nima PourNejatian, Hoo Chang Shin, Kaleb E Smith, Christopher Parisien, Colin Compas, Cheryl Martin, Mona G Flores, Ying Zhang, Tanja Magoc, Christopher A Harle, Gloria Lipori, Duane A Mitchell, William R Hogan, Elizabeth A Shenkman, Jiang Bian, Yonghui Wu

## Abstract

**Objective:** To develop a large pretrained clinical language model from scratch using transformer architecture; systematically examine how transformer models of different sizes could help 5 clinical natural language processing (NLP) tasks at different linguistic levels.

**Methods:** We created a large corpus with >90 billion words from clinical narratives (>82 billion words), scientific literature (6 billion words), and general English text (2.5 billion words). We developed GatorTron models from scratch using the BERT architecture of different sizes including 345 million, 3.9 billion, and 8.9 billion parameters, compared GatorTron with three existing transformer models in the clinical and biomedical domain on 5 different clinical NLP tasks including clinical concept extraction, relation extraction, semantic textual similarity, natural language inference, and medical question answering, to examine how large transformer models could help clinical NLP at different linguistic levels.

**Results and Conclusion:** GatorTron scaled up transformer-based clinical language models to a size of 8.9 billion parameters and achieved state-of-the-art performance on 5 clinical NLP tasks of different linguistic levels targeting various healthcare information documented in unstructured electronic health records (EHRs). The proposed GatorTron models performed remarkably better in much complex clinical NLP tasks such as natural language inference (9.6% and 7.5% improvements) and question answering (9.5% and 7.77% improvements) compared with existing smaller clinical transformer models (i.e., BioBERT and ClinicalBERT), demonstrating the potential of large transformer-based clinical models for advanced medical artificial intelligent (AI) applications such as question answering.

## INTRODUCTION

There has been an increasing interest in developing artificial intelligence (AI) systems by leveraging large electronic health records (EHRs). A critical step in developing medical AI systems is to enable machines to use patients’ clinical characteristics captured in longitudinal EHRs. The more information we know about patients, the better medical AI systems that we can develop. In recent decades, hospitals and medical practices in the United States (US) rapidly adopted EHR systems[1,2], resulting in massive stores of electronic patient data, including structured (e.g., disease codes, medication codes) and unstructured (i.e., clinical narratives such as physicians’ progress notes and discharge summaries). Physicians and other healthcare workers widely use clinical narratives to document detailed patient information as free text in EHRs. [3] There is an increasing number of studies exploring the rich, more fine-grained information about the patients in clinical narratives that led to improved diagnostic and prognostic models.[4,5] Nevertheless, free-text narratives cannot be easily used in computational models that usually require structured data. Researchers have increasingly turned to natural language processing (NLP) as the key technology to fill the gap of using clinical narratives in clinical studies[6].

Recently, transformer-based deep learning models have become state-of-the-art for many clinical NLP tasks. Compared with traditional machine learning models, transformer-based NLP models usually have very large number of parameters (e.g., 345 million parameters in BERT) to enable automated knowledge learning from a massive amount of text data. There is an increasing interest in examining how scaling up the model size could improve NLP. In the general NLP domain, many large transformer-based NLP models have been developed, such as the Generative Pre-trained Transformer 3 (GPT-3) model [7], which has 175 billion parameters and was trained using >400 billion words of text. However, few studies have explored large (i.e., billions of parameters) transformer models in the clinical domain. To date, the largest transformer model using clinical narratives, ClinicalBERT[8], has 110 million parameters and was trained using 0.5 billion words of clinical text. It is unclear how transformer-based models developed using significantly more clinical narrative text and more parameters may improve medical AI systems. In this study, we developed a large clinical transformer model, GatorTron, using >90 billion words of clinical narratives, scientific literature, and general English text. We trained GatorTron from scratch using UF’s HiperGator-AI cluster and empirically evaluated three models with different settings including (1) a base model with 345 million parameters, (2) a medium model with 3.9 billion parameters, and (3) a large model with 8.9 billion parameters. We compared GatorTron models with existing large transformer models trained using biomedical literature and clinical narratives on 5 clinical NLP tasks including clinical concept extraction (or clinical named entity recognition [CNER]), medical relation extraction (MRE), semantic textual similarity (STS), natural language inference (NLI), and medical question answering (MQA). GatorTron outperformed previous transformer models on 5 clinical NLP tasks at different linguistic levels targeting various patient information.

## BACKGROUND

Researchers have applied various methods including rule-based, machine learning-based, and hybrid solutions in clinical NLP. [9,10] At present, most state-of-the-art NLP models are based on machine learning models. For a long time, researchers had to train different machine learning models for different NLP tasks. Today, most state-of-the-art clinical NLP solutions are based on deep learning models[11] implemented using neural network architectures – a fast-developing sub-domain of machine learning. Convolutional neural networks[12] (CNN) and recurrent neural networks[13] (RNN) have been successfully applied to NLP in the early stage of deep learning. The RNN model implemented using bidirectional long-short term memory (LSTM) with a CRFs layer (LSTM-CRFs) has been widely used in CNER and relation extraction. [14–16] More recently, the transformer architectures[17] (e.g., BERT) implemented with a self-attention mechanism[18] have become state-of-the-art, achieving best performance on many NLP benchmarks. [19–22] In the general domain, the transformer-based NLP models have achieved state-of-the-art performance for many NLP tasks including name entity recognition[23–25], relation extraction[26–30], sentence similarity[31–33], natural language inference[33–36], and question answering[33,34,37,38]. Notably, transformers perform more effectively by decoupling of language model pretraining (i.e., pretrain language models using large unlabeled text corpora) and fine-tuning (i.e., applying the learned language models solving specific tasks often with labeled training data) into two independent phases. After successful pretraining, the learned language model can be used to solve a variety of NLP subtasks through fine-tuning, which is known as transfer learning – a strategy to learn knowledge from one task and apply it in another task[39]. Human language has a very large sample space – the possible combinations of words and sentences are innumerable. Recent studies show that large transformer models trained using massive text data are remarkably better than traditional NLP models in terms of emergence and homogenization.[39]

In the clinical domain, researchers have identified several fundamental NLP tasks such as CNER, MRE, STS, NLI, and MQA. CNER is to recognize phrases that have important clinical meanings (e.g., medications, treatments, adverse drug events). The NLP system has to determine the boundaries of a concept and classify it into predefined semantic categories. Early systems for clinical concept extract are often rule-based, yet, most recent systems are based on machine learning models such as conditional random fields (CRFs)[40,41], CNN [12,42], and LSTM-CRFs [13,14]. Current state-of-the-art solutions for CNER are mainly based on transformers[43]. MRE is to establish medical-related relations (e.g., drug induce adverse events) among clinical concepts (e.g., drugs, adverse events). MRE is usually approached as a classification problem – identify and classify pairs of concepts with valid relations. Various machine learning-based classifiers such as support vector machines (SVMs), random forests (RF), and gradient boosting trees (GBT)[16] have been applied. With the emergence of deep learning models, researchers have explored the LSTM architecture for RE in both general and clinical domains[44,45]. Most recently, several studies adopted the BERT architecture and demonstrated superior performance for MRE on various datasets[43,46–50]. The STS task is to quantitatively assess the semantic similarity between two text snippets (e.g., sentences), which is usually approached as a regression task where a real-value score was used to quantify the similarity between two text snippets. In the general domain, the STS benchmark (STS-B) dataset curated by the Semantic evaluation (SemEval) challenges between 2012 and 2017[51] is widely used for evaluating STS systems[19]. Various machine learning methods have been examined[52–54] but transformer-based systems such as RoBERTa[31], T5[33], and ALBERT[34] are leading the state-of-the-art models for STS. In the clinical domain, the MedSTS dataset[55] that consists of over 1,000 annotated sentence pairs from clinical notes at Mayo Clinic was widely used as the benchmark. MedSTS was used as the gold standard in two clinical NLP open challenges including the 2018 BioCreative/Open Health NLP (OHNLP) challenge[56] and 2019 n2c2/OHNLP ClinicalSTS shared task[57]. Similar to the general domain, pretrained transformer-based models using clinical text and biomedical literature, including ClinicalBERT and BioBERT[58], are current solutions for STS. NLI is also known as recognizing textual entailment (RTE) - a directional relation between text fragments (e.g., sentences)[59]. The goal of NLI is to determine if a given hypothesis can be inferred from a given premise. In the general domain, two benchmark datasets - the MultiNLI[60] and the Stanford NLI[61] are widely used. On both datasets, pretrained transformer models achieved state-of-the-art performances[33,35]. There are limited resources for NLI in the clinical domain. Until recently, the MedNLI – a dataset annotated by doctors based on the medical history of patients[62] was developed as a benchmark dataset in the clinical domain. A previous study[8] showed that a pretrained clinical BERT model achieved the state-of-the-art performance and outperformed the baseline (InferSent[63]) by ∼9% accuracy. The MQA task is to build NLP systems that automatically answer medical questions in a natural language. Unlike other tasks focusing on phrases and sentences, MQA is a document-level task that requires information from the whole document to generate answers according to questions. In the general domain, the Stanford Question Answering Datasets (SQuAD 1.1 and 2.0)[64,65] have been widely used as benchmarks. Transformer-based models are the state-of-the-art for both SQuAD1.1[24] and SQuAD2.0[37]. There are several MQA datasets developed in the past few years such as the MESHQA[66], MedQuAD[67], and emrQA[68].

The promise of transformer-based NLP models has led to further interest in exploring how increases in model and data size may improve large (e.g., >billions of parameters) transformer models processing clinical narratives. In the biomedical domain, researchers developed BioBERT[17] (with 110 million parameters) and PubMedBERT[69] (110 million parameters) transformer models using text from PubMed literature. Previously, we developed BioMegatron models in the biomedical domain with different sizes from 345 million to 1.2 billion parameters[70] using PubMed literature. However, few studies have explored large-size transformer models in the clinical domain due to the sensitive nature of clinical narratives that contain Protected Health Information (PHI) and the requirement of massive computing power. By developing not only larger models, but models that use clinical narratives, NLP may perform better in utilizing patient information in ways that can be applied to medical AI systems. To date, the largest transformer model using clinical narratives is ClinicalBERT[8]. ClinicalBERT has 110 million parameters and was trained using 0.5 billion words from the publicly available Medical Information Mart for Intensive Care III[71] (MIMIC-III) dataset. It is unclear how transformer-based models developed using significantly more clinical narrative text and more parameters may improve medical AI systems in extracting and utilizing patient information.

## MATERIALS AND METHODS

### Data Source

The primary data source for this study is the clinical narratives from UF Health Integrated Data Repository (IDR), a research data warehouse of UF Health. We collected a total of 290,482,002 clinical notes from 2011 to early 2021. The data included >82 billion medical words from >290 million notes related to >2 million patients and >50 million patient care encounters. We applied a standard preprocessing pipeline to remove duplicated notes and clean the clinical text – unify character encoding, identify tokens and sentence boundaries. Then, we merged the >82 billion words of clinical corpus with 6 billion words from PubMed (combining PubMed abstracts and full-text commercial-collection)[70], 2.5 billion words from Wikipedia[70], and 0.5 billion words from the MIMIC-III corpus[71] to generate a corpus with > 90 billion words. This study was approved by the UF Institutional Review Board (IRB202100049).

### Study design

**Figure 1** shows an overview of the study design. We seek to train a large clinical transformer model, GatorTron, using >90 billion words and examine how and whether scaling up mode size improves clinical NLP tasks. Following standard practice, we first pretrained GatorTron using the >90 billion words as an unsupervised learning procedure and then applied GatorTron to 5 different clinical NLP tasks using a supervised fine-tuning procedure. We adopted the BERT architecture implemented in MagaTron-LM[70] and explored three different settings including a base model of 345 million parameters (i.e., GatorTron-base), a medium model of 3.9 billion parameters (i.e., GatorTron-medium), and a large model of 8.9 billion parameters (i.e., GatorTron-large). Then we compared GatorTron models with an existing transformer model from the clinical domain, ClinicalBERT (trained with 110 million parameters) and two transformer models from the biomedical domain, including, BioBERT (345 million parameters) and BioMegatron (1.2 billion parameters). We examined the models on 5 clinical NLP tasks, including CNER, MRE, STS, NLI, and MQA. We used 6 public clinical benchmark datasets (**Table 2** and **Table 3**) following the default training/test settings and calculated evaluation scores using official evaluation scripts associated with each benchmark dataset.

**Figure 1.**
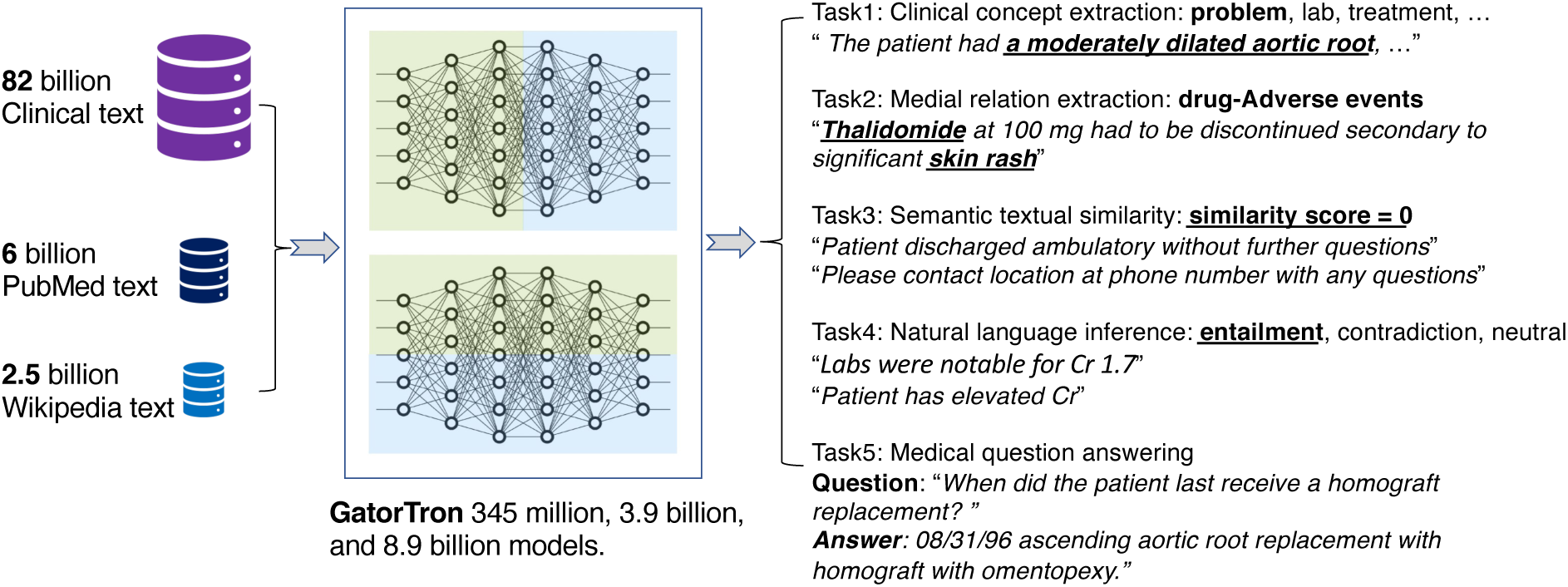
An overview of study design.

**Table 1.**
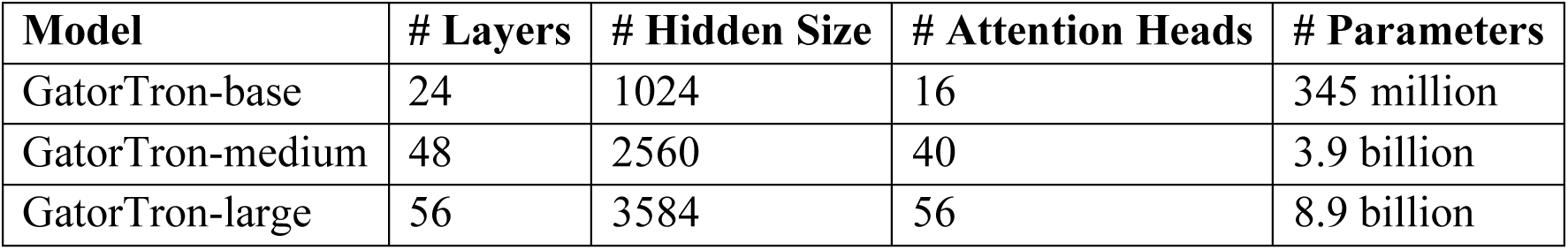
Three configurations of GatorTron model.

**Table 2.**
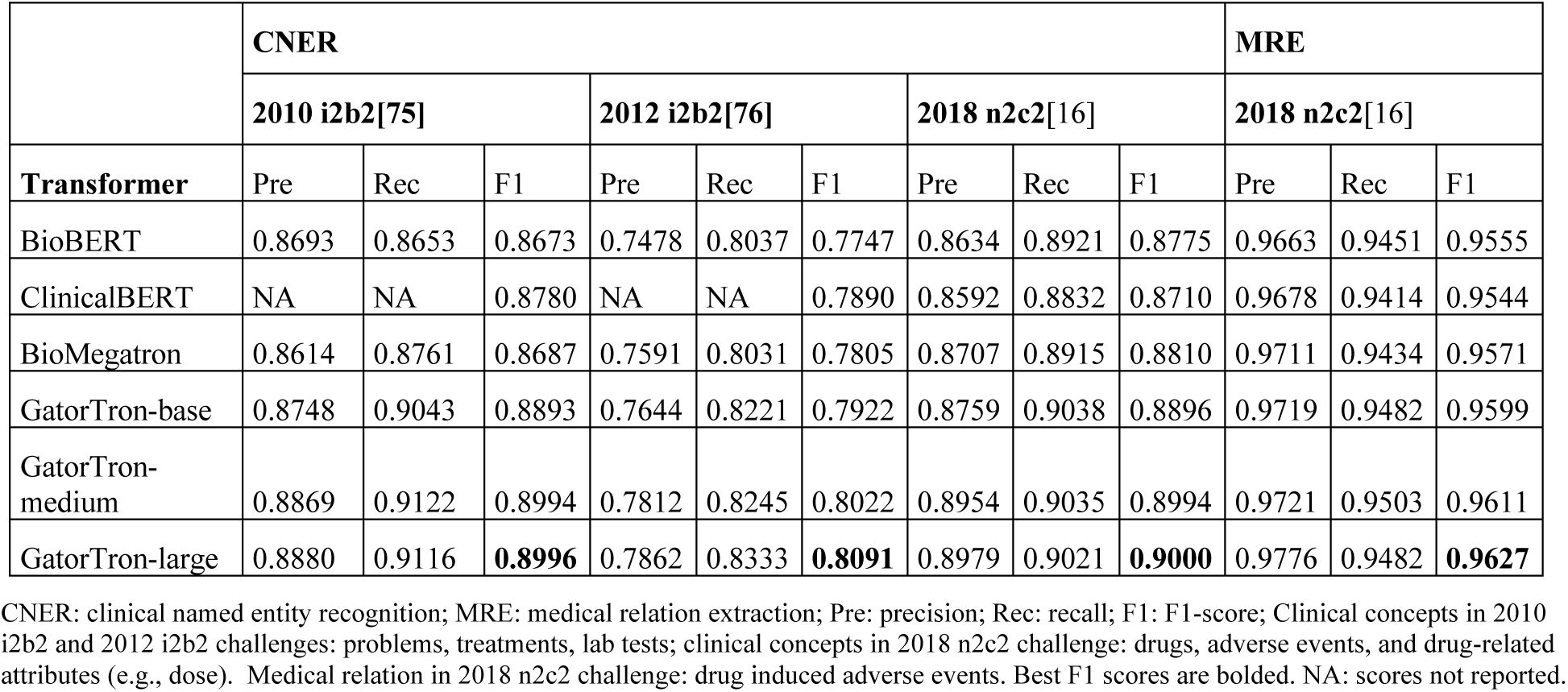
Comparison of GatorTron with existing biomedical and clinical transformer models for CNER and medical MRE.

**Table 3.** Comparison of GatorTron with existing biomedical and clinical transformer models for STS, NLI, and MQA.

|  | STS | NLI | MQA |  |  |  |
| --- | --- | --- | --- | --- | --- | --- |
|  | 2019 n2c2[57] | MedNLI[62] | emrQA Medication[68] |  | emrQA Relation[68] |  |
| Transformer | Pearson correlation | Accuracy | F1 score | Exact Match | F1 score | Exact Match |
| BioBERT | 0.8744 | 0.8050 | 0.6997 | 0.2475 | 0.9262 | 0.8361 |
| ClinicalBERT | 0.8787 | 0.8270 | 0.6905 | 0.2406 | 0.9306 | 0.8533 |
| BioMegatron | 0.8806 | 0.8390 | 0.7231 | 0.2882 | 0.9405 | 0.879 |
| GatorTron-base | 0.8810 | 0.8670 | 0.7181 | 0.2978 | 0.9543 | 0.9029 |
| GatorTron-medium | **0.8903** | 0.8720 | 0.7354 | 0.3018 | 0.9677 | 0.9243 |
| GatorTron-large | 0.8896 | **0.9020** | **0.7408** | **0.3155** | **0.9719** | **0.9310** |

STS: semantic textual similarity; NLI: natural language inference; MQA: medial question answering; The best evaluation scores are bolded.

### Training environment

We used a total number of 992 Nvidia DGX A100 GPUs from 124 superPOD nodes at UF’s HiperGator-AI cluster to train GatorTron models by leveraging both data-level and model-level parallelisms implemented by the Megatron-LM package[72]. We monitored the training progress by training loss and validation loss and stopped the training when there was no further improvement (i.e., the loss plot became flat).

### GatorTron Model Configuration

We developed GatorTron models using three configurations and determined the number of layers, hidden sizes, and number of attention heads according to the guidelines for optimal depth-to-width parameter allocation proposed by Levin et al[73] as well as our previous experience in developing BioMegatron[70]. **Table 1** provides detailed information for the three settings. The GatorTron-base model has 24 layers of transformer blocks, which is similar to the architecture of BERT large model. For each layer, we set the number of hidden units as 1024 and attention heads as 16. The GatorTron-medium model scaled up to 3.9 billion parameters (∼10 times of the base setting) and the GatorTron-large model scaled up to 8.9 billion parameters, which is similar to BioMegatron[72] (with 8.3 billion parameters).

### Existing transformer models for comparison

#### BioBERT

[17] The BioBERT model was developed by further training the original BERT-large model (345 million parameters, 24 layers, 1024 hidden units, and 16 attention heads) using biomedical literature from PubMed Abstracts (4.5 billion words) and PMC Full-text articles (13.5 billion words). In this study, we used version 1.1.

#### ClinicalBERT

[8] The ClinicalBERT model was developed by further training the BioBERT (base version; 110 million parameters with 12 layers, 768 hidden units, and 12 attention heads) using clinical text from the MIMIC-III[71] corpus.

#### BioMegatron

[70] The BioMegatron models adopted the BERT architecture with a different number of parameters from 345 million to 1.2 billion. Different from BioBERT and ClinicalBERT, the BioMegatron was trained from scratch without leveraging the original BERT model.

### Clinical NLP tasks, evaluation matrices, and benchmark datasets

We evaluated GatorTron models using 5 clinical NLP tasks and 6 public benchmark datasets. For CNER, we used three benchmark datasets developed by the 2010 i2b2 challenge, 2012 i2b2 challenge, and 2018 n2c2 challenge to evaluate GatorTron models on identifying various important medical concepts from clinical text. We used standard precision, recall, and F1-score for evaluation. For MRE, we used the dataset developed by the 2018 n2c2 challenge with a focus on relations between medications and adverse drug events. The standard precision, recall, and F1-score were used for evaluation. For STS, we used the dataset developed by the 2019 n2c2/OHNLP challenge on clinical semantic textural similarity[57]. We used the Pearson correlation score for evaluation. For NLI, we evaluated the Gatortron models using the MedNLI dataset and used accuracy for comparison. We used the emrQA dataset, a benchmark dataset widely used for MQA, to evaluate GatorTron. We particularly focused on medications and relations-related questions as Yue et al.[74] found that the two subsets are more consistent. We utilized both F1-score and exact match score for evaluation.

## RESULTS

**Table 2** and **Table 3** compare GatorTron models with two existing biomedical transformer models (BioBERT and BioMegatron) and one clinical transformer model (Clinical BERT) on 5 clinical NLP tasks.

### Recognize clinical concepts and medical relations

As shown in **Table 2**, all three GatorTron models outperformed existing biomedical and clinical transformer models in recognizing various types of clinical concepts on the three benchmark datasets (i.e., 2010 i2b2[75] and 2012 i2b2[76]: problem, treatments, lab tests; 2018 n2c2[16]: drug, adverse events, and drug-related attributes). The GatorTron-large model outperformed the other two smaller GatorTron models and achieved the best F1-scores of 0.8996, 0.8091, and 0.9000, respectively, demonstrating performance gain from scaling up the size of the model. For MRE, the GatorTron-large model also achieved the best F1-score of 0.9627 for identifying drug-cause-adverse event relations outperforming existing biomedical and clinical transformers and the other two smaller GatorTron models. We observed performance improvement when scaling up the size of GatorTron model.

### Assess semantic textual similarity

As shown in **Table 3**, all GatorTron models outperformed existing biomedical and clinical transformer models in assessing STS. Among the three GatorTron models, the GatorTron-medium model achieved the best Pearson correlation score of 0.8903, outperforming both GatorTron-base and GatorTron-large. Although we did not observe consistent improvement by scaling up the size of the GatorTron model, the GatorTron-large model significantly outperformed GatorTron-base and its performance is very close to the GatorTron-medium model (0.8896 vs. 0.8903).

### Natural language inference

GatorTron models outperformed existing biomedical and clinical transformers, and the GatorTron-large model achieved the best accuracy of 0.9020, outperforming the BioBERT and ClinicalBERT by 9.6% and 7.5%, respectively. We observed a monotonic performance improvement by scaling up the size of GatorTron.

### Medical question answering

All GatorTron models outperformed existing biomedical and clinical transformer models in MQA (e.g., “What lab results does patient have that are pertinent to diabetes diagnosis?”). For medication-related questions, the GatorTron-large model achieved the best exact match score of 0.3155, outperforming the BioBERT and ClinicalBERT by 6.8% and 7.5%, respectively. For relation-related questions, GatorTron-large also achieved the best exact match score of 0.9301, outperforming BioBERT and ClinicalBERT by 9.5% and 7.77%, respectively. We also observed a monotonic performance improvement by scaling up the model size of GatorTron.

## DISCUSSION

In this study, we developed a large pretrained language model, GatorTron, using a corpus of >90 billion words. We trained GatorTron from scratch with different model sizes and evaluated its performance on 5 clinical NLP tasks at different linguistic levels (phrase level, sentence level, and document level) using 6 publicly-available benchmark datasets from the clinical domain. The experimental results show that GatorTron models outperformed existing biomedical and clinical transformers for all 5 clinical NLP tasks. We observed monotonic improvements by scaling up the model size of GatorTron for 4 of the 5 tasks, excluding the STS task. Our GatorTron model also outperformed the BioMegatron[70], a transformer model with a similar model size developed in our previous study using >8.5 billion words from PubMed and Wikipedia (a small proportion of the >90 billion words of corpus for developing GatorTron). This study scaled up the clinical transformer models to 8.9 billion parameters in the clinical domain and demonstrated performance improvements. To the best of our knowledge, GatorTron-large is the largest transformer model in the clinical domain.

### Scaling up model size and performance improvement

There is an increasing interest in examining massive-size deep learning models in NLP as they demonstrated novel abilities such as emergence and homogenization[39]. In the general domain, the Megatron-Turing NLG model has scaled up to 530 billion parameters following the GPT-3[7] model with 175 billion parameters. However, there are limited studies examining large transformer models in the clinical domain due to the sensitive nature of clinical text and massive computing requirements. Prior to our study, the largest transformer in the clinical domain was ClinicalBERT with 110 million parameters trained using 0.5 billion words. Our study scaled the transformer to 8.9 billion parameters and demonstrated performance improvement for 5 clinical NLP tasks on 6 public benchmark datasets. Among the 5 tasks, GatorTron achieved significant improvements for sentence-level and document-level NLP tasks such as NLI and MQA, but moderate improvements for phrase-level tasks such as CNER and MRE, indicating that large transformer models are more helpful to sentence-level and document-level NLP tasks.

### Model size and converge speed

GatorTron was pretrained using unsupervised learning to optimize a mask language model (MLM). We monitored training loss and calculated validation loss using a subset set of the clinical text (5%) to determine when to stop the training. **Figure 2** shows the training loss and validation loss for GatorTron models with three different settings.

**Figure 2.**
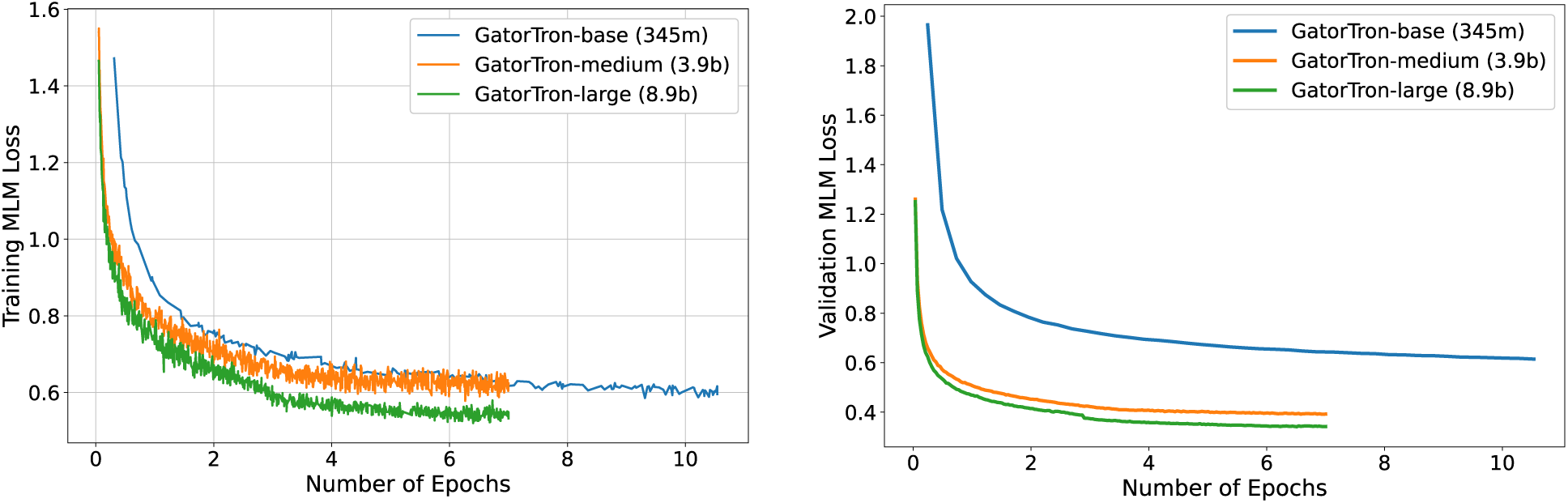
Training loss and validation loss for GatorTron base (345 million), medium (3.9 billion), and large (8.9 billion) models.

We observed that the larger GatorTron models converged faster than the base model. For example, the GatorTron-base model converged in 10 epochs, whereas the medium and large models converged in 7 epochs. This may indicate that larger transformer models learn faster than smaller models. The training of the GatorTron-large model used about 6 days on 992 GPUs from 124 Nvidia SuperPOD nodes.

### Potentials in improving healthcare delivery and patient outcomes

GatorTron models perform better in utilizing patient information in clinical narratives, which can be applied to various medical AI systems. The rich, fine-grained patients’ information captured in clinical narratives is a critical resource powering medical AI system. With better performance in information extraction tasks (e.g., CNER and MRE), GatorTron models have potential to provide more accurate patients’ information to identify research-standard patient cohorts using computable phenotypes, support physicians making data-informed decisions by clinical decision support systems, and identify adverse events associated with drug exposures via pharmacovigilance. The significant improvements in STS, NLI, and MQA can be applied for deduplication of clinical text, mining medial knowledge, and developing next-generation medical AI systems that can interact with patients using human language. The emergence and homogenization abilities[39] inherited from a large transformer architecture make it convenient to apply GatorTron to many other AI tasks through fine-tuning. We believe that GatorTron will improve the use of clinical narratives in developing various medical AI systems for better healthcare delivery and health outcomes.

This study has limitations. We mainly focused on medication and relation-related questions when evaluating GatorTron models due to the limitation of the benchmark dataset for MQA. Future studies should examine the benefit of large clinical transformer models to downstream medical applications such as disease phenotyping and patient cohort construction.

## CONCLUSION

Large pretrained clinical language models could benefit a number of downstream clinical NLP tasks, especially for complex NLP tasks such as MQA.

## Data Availability

All data produced are available online at N2C2 https://n2c2.dbmi.hms.harvard.edu/data-sets

## ACKNOWLEDGMENTS

None

## FUNDING STATEMENT

This study was partially supported by a Patient-Centered Outcomes Research Institute® (PCORI®) Award (ME-2018C3-14754), a grant from the National Cancer Institute, 1R01CA246418 R01, grants from the National Institute on Aging, NIA R56AG069880 and R21AG062884, and the Cancer Informatics and eHealth core jointly supported by the UF Health Cancer Center and the UF Clinical and Translational Science Institute. The content is solely the responsibility of the authors and does not necessarily represent the official views of the funding institutions.

### Support from UF Research Computing

We would like to thank the UF Research Computing team, led by Dr. Erik Deumens, for providing computing power through UF HiperGator-AI cluster.

## CONTRIBUTORSHIP STATEMENT

XY, YW, JB, NP, and MGF were responsible for the overall design, development, and evaluation of this study. XY had full access to all the data in the study, conducted all the experiments, and takes responsibility for the integrity of the data and the accuracy of the data analysis. XY, YW, JB, and WH did the bulk of the writing, EAS, DAM, TM and CAH also contributed to writing and editing of this manuscript. All authors reviewed the manuscript critically for scientific content, and all authors gave final approval of the manuscript for publication.

## ETHICS STATEMENT

IRB (202100049) of the University of Florida gave approval for this work as exempt. The approval includes but is not limited to HIPAA waiver to enroll.

## SUPPLEMENTARY MATERIAL

None.

